# Sterilizable, Time-Integrating Hydrogel Sensors Enable Continuous Gastrointestinal Leak Surveillance in Low- and High-Resource Settings

**DOI:** 10.64898/2026.07.21.26358572

**Authors:** Alexander Jessernig, Isabella von Forcade de Biaix, Carolin Himmel, Sergio Alejandro Gómez-Ochoa, Annina Wolf, Felix Spengler, Juan Carlos Hernandez-Vargas, Diana Clemencia Quintero-Gamboa, Javier Mauricio Pacheco-Maldonado, Juan Paulo Serrano-Pastrana, Andrea Schlegel, Andrea Carolina Quiroga-Centeno, Ignazio Tarantino, Inge K. Herrmann

## Abstract

Gastrointestinal anastomotic leakage (AL) remains a life-threatening complication following gastrointestinal surgery, where outcomes critically depend on timely diagnosis. Current diagnostic strategies rely on periodic sampling and resource-intensive analysis in centralized laboratories.

Here, we present a sterilizable, time-integrating hydrogel sensor platform for continuous, infrastructure-free monitoring of the patient’s postoperative drain fluid. We introduce enzyme-responsive macromolecular networks for semi-quantitative bedside assessment of leak-associated digestive enzymes. The sensors retain functionality following lyophilization and ethylene oxide sterilization, enabling long-term storage and scalable deployment around the world. In a Swiss clinical cohort of 56 patients, including 19 with gastrointestinal anastomotic leakage, the sensor detected amylase-associated leaks two days (median) prior to clinical diagnosis with a sensitivity of 78% (95% CI 55-91) and a specificity of 95% (95% CI 82-99). The prospective validation in an independent cohort of 37 patients in Colombia, including seven patients with leaks, demonstrated 100% sensitivity (95% CI 64.6-100) and a 100% negative predictive value (95% CI 87.9-100.0), with sensor activation preceding standard clinical diagnosis by a median of five days. By converting episodic biochemical measurements into continuous, cumulative visual records, this infrastructure-free material platform enables close-meshed postoperative monitoring and may facilitate earlier recognition of anastomotic leakage across diverse healthcare settings.

## INTRODUCTION

Each year, an estimated 14 million individuals worldwide undergo abdominal surgery,^1^ with gastrointestinal (GI) anastomoses routinely performed to restore bowel continuity after resection of diseased segments.^2^ While these procedures are often life-saving and essential for maintaining quality of life, they expose patients to the risk of gastrointestinal anastomotic leak (AL), a serious postoperative complication. AL incidence rates fluctuate based on surgical location,^3–5^ technical aspects of the procedure, as well as the patients’ predisposition, such as malnutrition, cardiovascular diseases, patient lifestyle (alcohol, smoking etc.), and preoperative chemotherapy.^6,7^ Additionally, the number of interventions at risk of AL is expected to rise significantly in the coming years, with e.g. cancer-related GI surgery projected to increase by over 53% by 2040, particularly in low- and middle-income countries.^8^

Among gastrointestinal procedures, major pancreatic resections represent a particularly high-risk setting with reported leak rates varying between 3-45% and mortality rates of up to 25% in the case of grade C-Postoperative pancreatic fistula.^9,10^ The high mortality is largely attributable to delayed diagnosis, which may lead to diffuse peritonitis, sepsis and potentially secondary organ dysfunction as well as life-threatening hemorrhage caused by erosion of adjacent blood vessels.^11–13^ Despite the substantial morbidity and mortality associated with anastomotic leaks, current diagnostic approaches remain insufficient for reliable early detection, with diagnosis typically occurring 5-8 days after surgery.^14^

Current clinical practice primarily relies on clinical assessment of the patient’s overall status and on monitoring systemic inflammatory biomarkers, such as C-reactive protein (CRP) and leukocyte count.^15^ When surgical drains are left in place, qualitative assessment of the drain fluid, including color, turbidity, odor, and related features, may also contribute to clinical evaluation, and, in some settings, this may be complemented by measurement of enzymatic biomarkers such as amylase in centralized lab facilities.^16^ While systemic inflammatory markers, particularly when combined with computed tomography (CT), may provide good negative predictive value, they are often nonspecific, lack sensitivity for detecting small leaks, and expose patients to harmful ionizing radiation.^17,18^ In contrast, drain amylase is a well-established marker of pancreatic leaks and has also shown predictive value for leaks at other sites in the gastrointestinal tract, irrespective of patient comorbidities.^19–24^ A shared limitation of these approaches is their discontinuousness, confining leak status assessment to isolated discrete measurements rather than continuous monitoring. Furthermore, they require specialized laboratory infrastructure and trained personnel. These constraints intrinsically delay diagnosis, forcing reliance on nonspecific, often late clinical symptoms such as pain or fever before additional testing is initiated.

In recent years, researchers have explored the development of technologies aimed at improving patient outcomes through continuous monitoring of anastomoses. Most of these approaches rely on measuring pH changes in biofluids, often coupled with AI-based data processing, through either electrodes positioned at the surgical site (e.g., Exero Medical)^25^ or external sensing devices incorporated into the drain line between the patient and the collection reservoir (e.g. FluidAI Medical).^26^ While these systems have demonstrated feasibility in selected clinical settings, their broader clinical utility remains limited. pH reflects a global metabolic parameter and can be influenced by a range of physiological and pathological processes, including inflammation, ischemia, and infection, thereby limiting its specificity for the detection of anastomotic leaks.^27^ Furthermore pH based methods require frequent calibration and, in some cases invasive components. Another major drawback is their reliance on electronic systems, which are typically associated with substantial acquisition costs and ongoing operational and maintenance expenses for hospitals.^28^ In view of the limitations of pH-based monitoring, enzyme-based electronic detection strategies have been explored more recently.^29^ Given the pathophysiological relevance and catalytic amplification properties of digestive enzymes as biomarkers, we recently demonstrated the development and characterization of a non-electronic, enzyme-responsive macromolecular network sensor.^30^ While enzyme-responsive sensing approaches may offer improved specificity compared to pH-based systems, these have not yet been validated using clinical patient cohorts. Collectively, these limitations highlight the need for a detection strategy that combines specificity with simplicity, while avoiding reliance on complex electronic systems or mandatory laboratory capacity.

Here, we report the design of a fully field-compatible clinically relevant hydrogel sensing array along with real-world validation in two distinctly different hospital settings (Figure 1a). The sensor array consisting of three enzyme-responsive hydrogels can be readily integrated into standard surgical drain bags (Figure 1b). In the event of an anastomotic leak, the sensing elements respond to the presence of digestive enzymes, thereby providing a direct and easily interpretable visual alert to clinicians. The sensor performance was validated in two independent clinical cohorts, a study comprising 56 patients in Switzerland, where patient samples were tested to assess the sensor’s performance, and a prospective cohort with on-site evaluation in 37 patients in Colombia, under real-life conditions in a resource-limited hospital environment.

**Figure 1:**
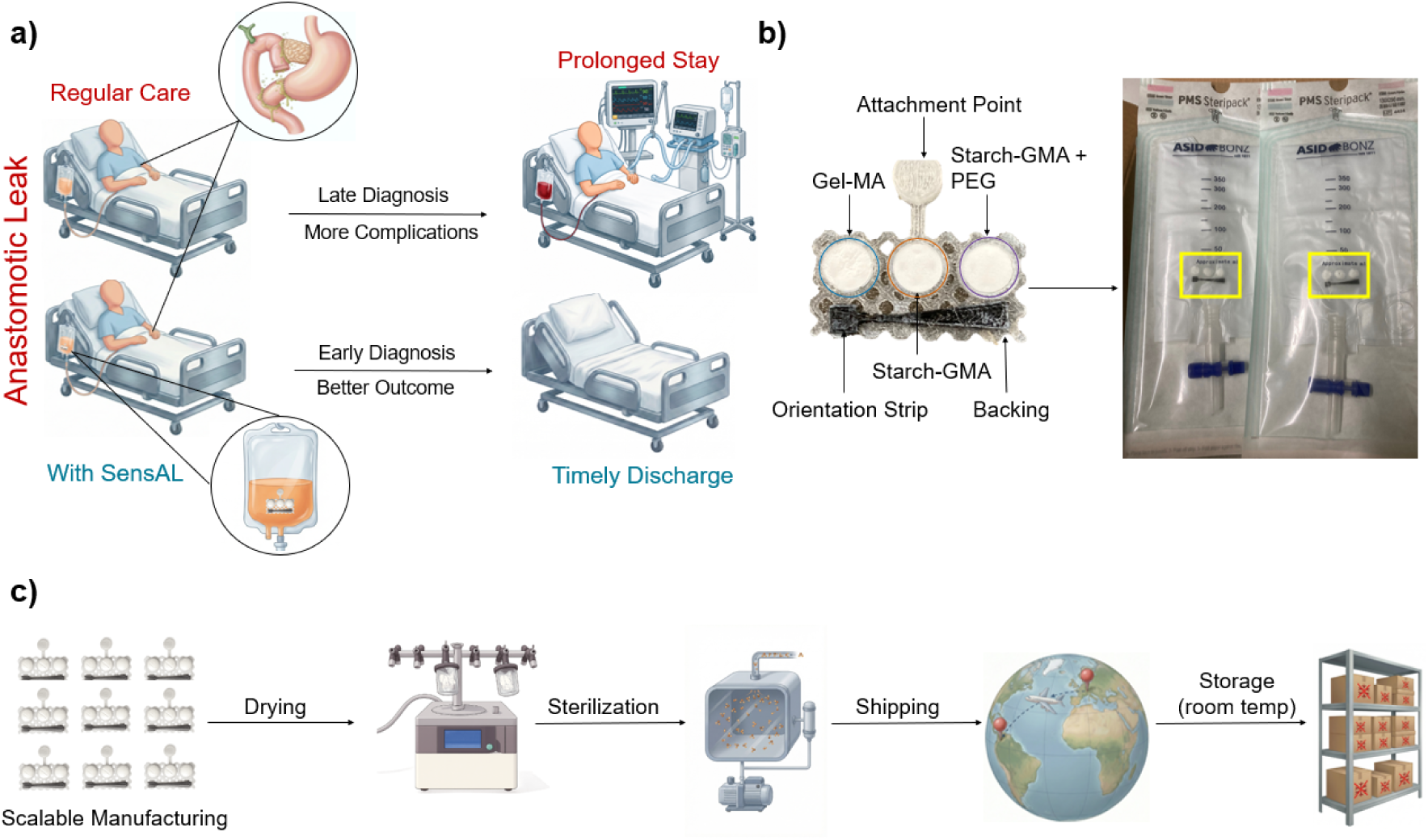
Schematic overview of the working principle and manufacturing of SensAL. a) SensAL can be easily integrated into regular standard of care. In the case of a leak, digestive enzymes in the fluid react with the sensor causing sensing element discoloration. b) The sensor contains a trypsin responsive sensing element as well as two amylase responsive sensing elements encased in a 3D printed backing. The sensor array can be readily integrated into drain fluid bags. c) Sensor manufacturing process of SensAL. Freeze-drying in combination with subsequent sterilization allows the sensor to be easily shipped and stored under ambient conditions until being used.

## RESULTS & DISCUSSION

### Sensing Element Design, Synthesis and Characterization

Since high digestive enzyme concentrations are normally limited to the gastrointestinal tract, elevated levels in abdominal drain fluid are indicative of intraluminal contents spillage and may support the diagnosis of an anastomotic leak (AL).^20^ On this basis, we designed enzyme-responsive hydrogel sensors (SensAL) for the sensitive and selective detection of digestive enzymes in postoperative drain fluid.^30^ These sensing elements contain enzyme-specific macromolecular substrates and a coloring agent (here TiO_2_ particles, a white pigment, enabling optimal visibility in drain fluids) integrated into the crosslinked hydrogel networks. Upon exposure to the corresponding digestive enzyme, the network undergoes selective enzymatic degradation, resulting in the breakdown of the macromolecular structure. This degradation process and the associated change in structure translates enzymatic activity in postoperative drain fluid into a naked-eye detectable response.

As the response is governed by enzyme-specific cleavage of the polymer networks, high selectivity toward digestive enzymes is achieved.^30^ Furthermore, by tailoring the crosslink density and network composition, the rate and extent of degradation can be tuned, enabling detection within clinically relevant enzyme concentration ranges and allowing for semi-quantitative readout of enzymatic activity in postoperative drain fluid. For clinical translation, several material and functional requirements must be met, including robust performance under continuous exposure to complex biological fluids, as well as the ability to tune sensor response across clinically relevant enzyme concentration ranges. Importantly, the enzyme-responsive hydrogel sensors need to retain their structural integrity and functional responsiveness following lyophilization and sterilization, enabling long-term storage and facilitating scalable, regulation-compliant manufacturing and deployment in clinical settings.

Starch-based hydrogel systems are attractive candidates for amylase-responsive sensing due to their biocompatibility and substrate specificity. However, conventional starch-based hydrogels often lack sufficient functionalization to form stable, independently crosslinked networks, necessitating the incorporation of additional synthetic polymers to achieve structural integrity.^30^ To overcome these limitations and to incorporate adjustable crosslinking strategies for tunable degradation behavior, starch was functionalized in dimethyl sulfoxide (DMSO) using glycidyl methacrylate (GMA). DMSO promotes enhanced swelling of otherwise water-insoluble starch granules,^31^ improving reaction homogeneity, while GMA facilitates efficient methacrylate incorporation through epoxide ring opening, offering higher functionalization efficiency compared to transesterification with methacrylic anhydride (see Figure 2a).^32^ This increased degree of functionalization enables precise control over crosslink density, thereby facilitating tunable degradation behavior and enabling semi-quantitative readout for amylase-based detection. The granular morphology of the starch is shown in Figure 2b (top left). During functionalization, the micrometer-sized crystalline starch particles swell and elongate substantially but do not fully dissolve, as indicated by filament-like structures with bulbous termini (top middle). After polymerization into a hydrogel, these structures remain discernible, forming a heterogeneous network in which elongated modified starch strands are interconnected through crosslinking (top right). In contrast, native gelatin possesses a more amorphous morphology and larger particle size (bottom left). When dissolved in water at elevated temperatures, gelatin fully solubilizes during the reaction process. Upon cooling and drying, Gel-MA forms a percolated network of interconnected biopolymer chains (bottom middle), and subsequent polymerization results in a comparatively homogeneous hydrogel structure (bottom right). This functionalization strategy enables improved crosslinking of Starch-GMA, allowing degradation to be tuned to clinically relevant thresholds which is shown in the degradation kinetics of the Starch-GMA sensing elements (Figure 3a). Specifically, the sensing elements respond to extremely high amylase activities (100,000 U/L also found within one patient in this study) within 5 hours, and to concentrations associated with typical clinically significant grade B or C pancreatic fistulas (amylase activities exceeding 5,000 U/L), which require active intervention such as reoperation, within approximately 12 hours.^10,33^ The sensor further detected amylase activity at the lowest threshold considered clinically indicative of a leak (≥1000 U/L)^34,35^ within 30 hours. Importantly, these detection times are approximately twice as fast as the interval between periodic drain-fluid measurements in hospitals equipped to perform such analyses. The sensing elements remained stable below this threshold, underscoring their ability to distinguish leak-relevant enzyme activity from background levels. Notably, comparable degradation behavior was observed for sterilized and lyophilized sensing elements, indicating that these processing steps do not compromise sensor performance (see also Figure 3b for the digestion kinetics of native and sterilized GelMA). In addition, incorporation of polyethylene glycol diacrylate (PEGDA) as a secondary crosslinker provides further control over the hydrogel network architecture. By incorporating PEGDA into the network, sensing elements remain stable under moderate postoperative amylase elevations (e.g., ∼100 U/L) that may occur in the absence of a true leak, thereby eliminating false-positive readouts from sustained background enzyme activity. However, in the event of a true leak, characterized by a rapid and sustained rise in amylase activity, the PEGDA-reinforced elements degrade alongside the non-reinforced sensing elements, providing confirmatory evidence of clinically significant leakage.

**Figure 2:**
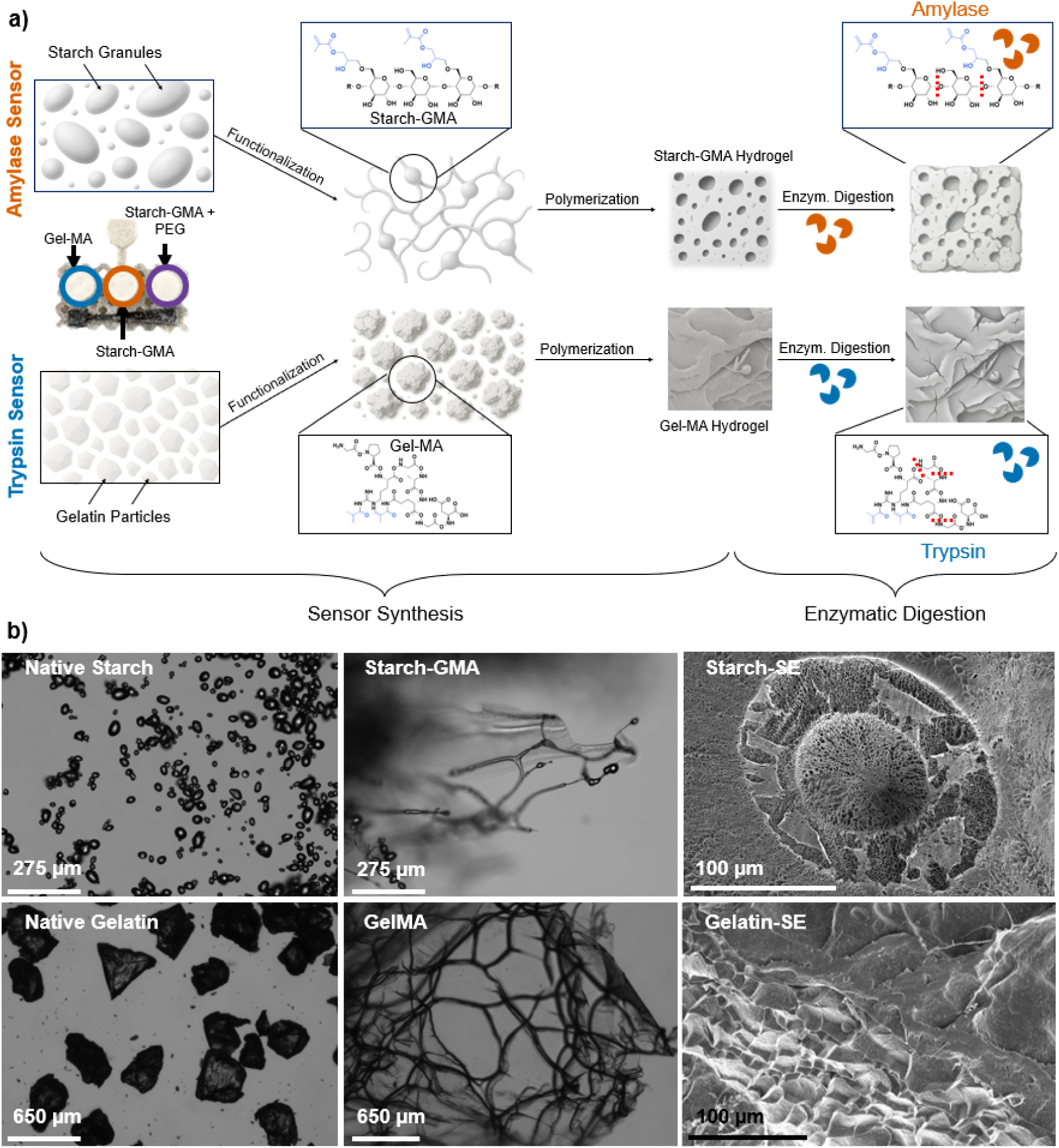
Synthesis and functionalization of SensAL. a) Schematic overview of the chemical functionalization and degradation of the biopolymer-based sensing elements. b) The top row shows light microscopy images of the native starch granules before (left) and after (middle) functionalization with GMA. Through the functionalization the granules morph into strings that then form a heterogeneous hydrogel after polymerization (right, SEM). The bottom row shows the native gelatin particles before (left) and after (middle) functionalization with MA. Gelatin is fully soluble in water and thus Gel-MA forms homogenous hydrogels (right).

**Figure 3:**
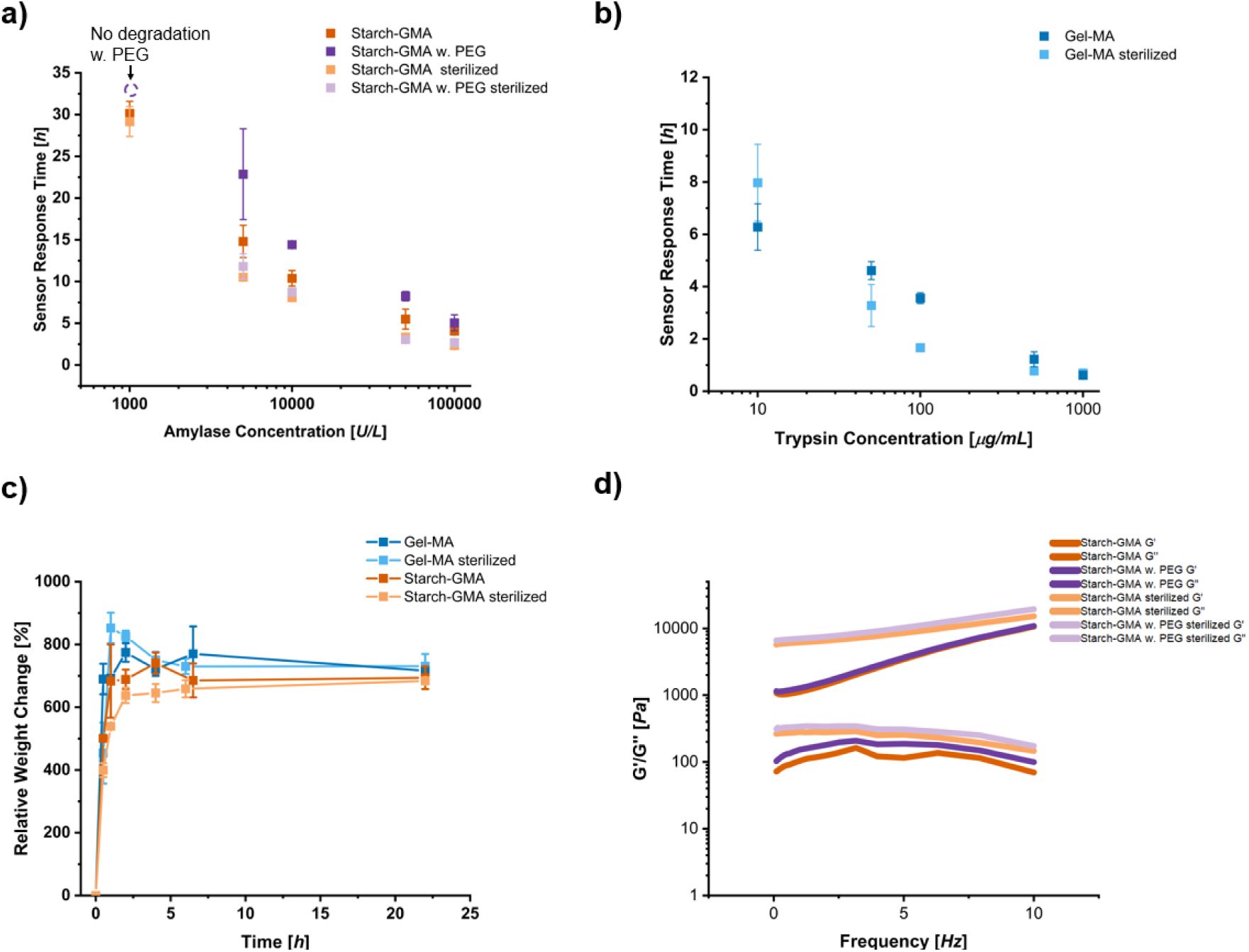
Characterization of the sensing elements. a) Digestion kinetics of freshly prepared hydrated hydrogel sensors, and sterilized and stored sensing elements when exposed to defined amylase concentrations. b) Digestion kinetics of freshly prepared (as previously published^30^) and sterilized GelMA sensing elements when exposed to Trypsin. Lyophilization and sterilization show little impact on digestion kinetics. c) Rheological measurements of Starch-GMA before and after sterilization. Sterilization or addition of PEG has only a limited effect on mechanical behavior. d) Swelling behavior of native starch and gelatin hydrogels before and after sterilization. Both lyophilized and sterilized hydrogels reach a swelling equilibrium after roughly 5h while having superior storage properties compared to hydrated hydrogels.

By combining readily available biopolymers with a simple one-step synthesis and a scalable design, the sensor can potentially be manufactured at costs well below 1 USD per unit. Accordingly, we sought to develop a system that is not only inexpensive to produce but also easy to store and handle, enabling stable storage at room temperature without specialized infrastructure, an important consideration for deployment in low- and middle-income countries. The preparation process is illustrated in Figure 1c. Briefly, freshly polymerized sensors are lyophilized, incorporated into commercially available surgical drainage bags, and subsequently sterilized to yield a ready-to-use device suitable for shipping and hospital storage. To ensure that this fabrication process does not alter the sensor’s rapid responsiveness and that they remain securely positioned within their backing structure, we compared the swelling behavior and rheological properties of freshly prepared hydrated sensing elements with those of fully processed, ready-to-use sensors. Figure 3c presents the swelling behavior of processed and native Starch-GMA and Gel-MA hydrogels over a 24 h period. No significant differences were observed between the processed and native gels, with both hydrogel systems reaching equilibrium swelling after approximately 5 h. Similarly, Figure 3d shows the frequency-dependent viscoelastic properties of Starch-GMA and Starch-GMA/PEG hydrogels, native and after lyophilization and rehydration. Across all conditions, G′ consistently exceeds G″ by approximately one order of magnitude over the 0–10 Hz range, confirming a predominantly elastic, solid-like network characteristic of chemically crosslinked hydrogels. PEG-containing formulations display substantially higher moduli (∼5,000–10,000 Pa versus ∼1,000 Pa for Starch-GMA alone), indicating that PEG incorporation significantly reinforces network stiffness. Following lyophilization and rehydration, both G′ and G″ were approximately 5-fold higher compared to native hydrogels, suggesting structural reorganization during freeze-drying that persists upon rehydration. Contrary to PEG incorporation, which also resulted in increased network stiffness, the lyophilization-induced stiffening did not translate into any measurable change in sensor responsiveness, as confirmed by the digestion kinetics in Figure 3a.

The optimized sensing element chemistry and sensor response times pave the way to highly sensitive detection of leak events while maintaining stability in the absence of leaks, even during prolonged exposure to postoperative drain fluid. Importantly, the fabrication process preserves the sensor’s functional properties while enabling extended shelf life and simple storage conditions, features that are particularly advantageous for deployment in resource-limited settings.

### Sensor Performance and Validation

To evaluate the sensor’s performance in a clinical context, drain fluid samples from 56 patients undergoing surgery (predominantly pancreatic resection; patient information in additional SI file) were collected on each postoperative day (POD). Among these, 19 patients (33.9%; 95% CI 22.9–47.0) were diagnosed with a clinically evident leak, as diagnosed by hospital standard (for further information see patient data file in the SI). Each sample was analyzed in a blinded fashion with both the hydrogel sensor array (SensAL) and the clinical reference standard, consisting of certified clinical-chemistry amylase activity. Overall amylase concentration was markedly higher in the leak group compared to the non-leak group (p = 4×10 ¹²), with the most pronounced separation observed in the early postoperative period and this difference progressively diminishing over time (p = 0.004) (see Figure 4a and b, for specific values see patient data tables in the SI). On POD1, the model-estimated geometric-mean amylase concentration was approximately 28-fold higher in leak than in no-leak patients (3,172 versus 115 U/L; adjusted fold-change 27.5, 95% CI 12.8–59.0), and the difference remained substantial on POD3 (513 versus 28 U/L; 18.2-fold, 95% CI 8.0–41.3) (see Figure 4a and b).

**Figure 4.**
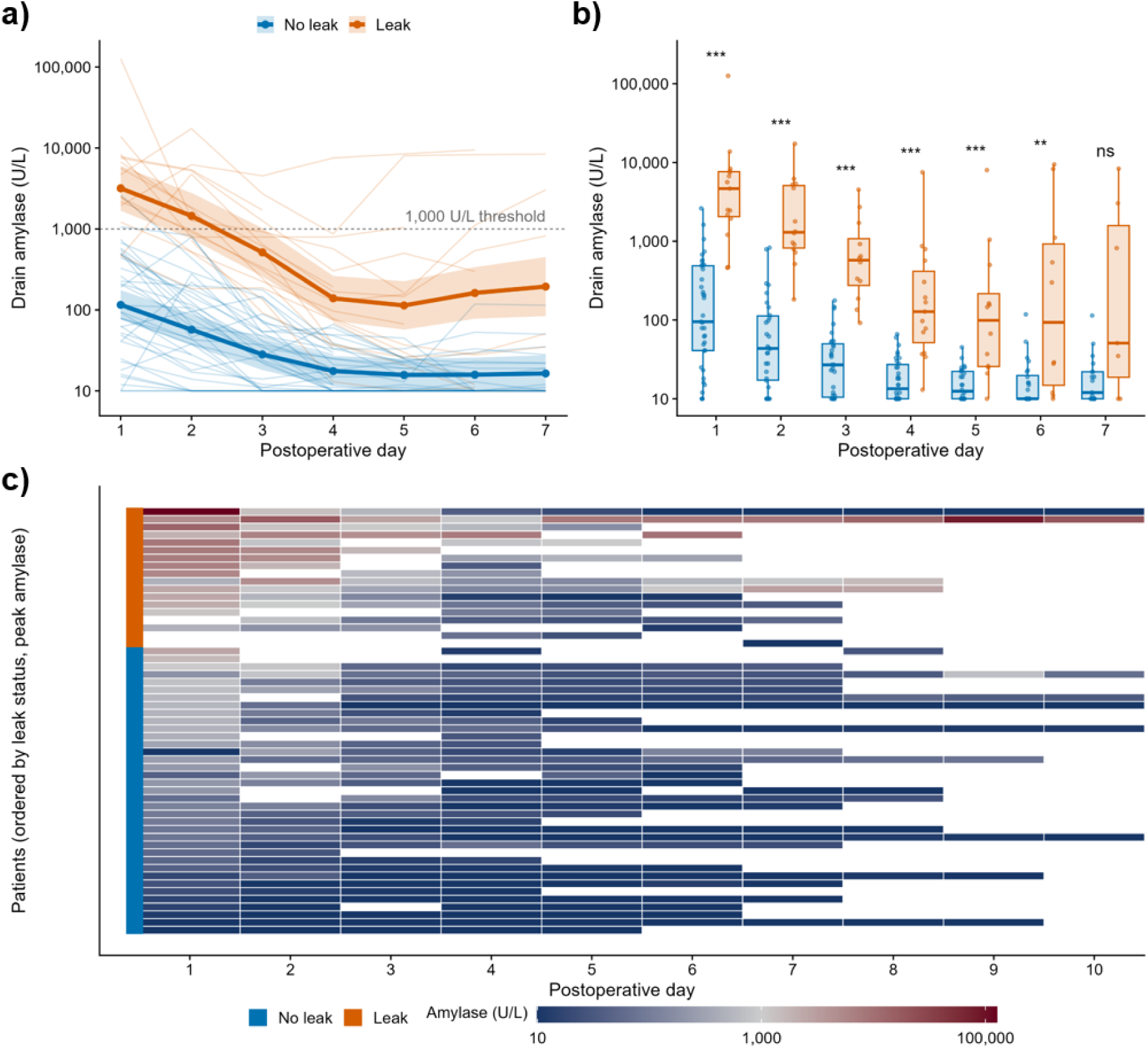
Postoperative drain amylase dynamics (Switzerland cohort). a) Individual patient amylase trajectories (thin lines) with linear mixed-model geometric means and 95% confidence intervals (bold lines) on a log scale; the dashed line marks the 1,000 U/L sensor threshold. b) Drain amylase by postoperative day in leak versus non-leak patients (box-and-jitter, log scale); asterisks denote Benjamini–Hochberg-adjusted Wilcoxon significance (*** q < 0.001, ** q < 0.01, * q < 0.05; ns, not significant), significant on POD1–6 (POD7 ns). c) Heatmap of drain amylase across all patients (rows, ordered by leak status then peak amylase) and postoperative days; the colored band on the left indicates leak status.

However, the reduced separation observed at later postoperative time points likely reflects not a biological phenomenon, but rather a consequence of clinical recognition and the therapeutic interventions initiated around POD3. Notably, a clearly elevated biochemical signal is already present early after surgery, as illustrated in Figure 4c. This suggests that surveillance initiated immediately postoperatively could allow leaks to be identified at an early stage and, importantly, could provide information not only on the presence of a leak but also on its evolution over time. Such continuous monitoring may help clinicians make earlier decisions regarding the need for additional interventions. In contrast, frequent biochemical analysis of drain fluid can be logistically challenging and financially burdensome. The main advantage of SensAL is its bedside applicability, enabling real-time leak detection and longitudinal tracking without the logistical burden of repeated laboratory measurements.

To quantify the diagnostic potential of this early biochemical signal, ROC analysis was performed on the measured amylase concentrations to evaluate their ability to discriminate between leak and no-leak patients. ROC curves generated using the maximum amylase value recorded per patient across all postoperative days, as well as separately for POD1 and POD3 (Figure 5a-c), demonstrated strong discriminative performance between leak and non-leak patients. For the maximum amylase concentration per patient, the analysis yielded an AUC of 0.88 (95% CI 0.75-1.00), while considering individual postoperative days further improved performance, with AUC values of 0.95 (95% CI 0.90-1.00) for POD1 and 0.98 (95% CI 0.96-1.00) for POD3 (Figure 5a-c). These findings are in line with previously reported AUC values and confirm that elevated drain amylase provides a robust biomarker for identifying pancreatic anastomotic leaks,^36^ with particularly strong discriminative power in the early postoperative period. Having established that drain amylase levels discriminate reliably between leak and no-leak patients, we next evaluated whether SensAL could translate this biochemical signal into a direct visual readout. Sensor responses to the collected drain fluid samples were assessed at each POD using time-lapse imaging over 48 hours. A sensor response was defined as positive when complete enzyme-induced degradation of a sensing element was observed. At the patient level, a case was classified as true positive (TP) when at least one positive sensor response occurred across the monitored postoperative days (PODs) in a patient clinically diagnosed with an anastomotic leak. As the sensor responds to amylase, one patient with an isolated bile leak (in whom amylase remained at 20 U/L, consistent with the absence of pancreatic enzyme in bile) lies outside the detection scope of an amylase sensor and was therefore not included. On this basis, the sensor correctly identified 14 of 18 leaks and correctly remained negative in 35 of 37 patients without a leak, corresponding to a sensitivity of 78% (95% CI 55-91) and a specificity of 95% (95% CI 82-99) (Figure 5d). Notably, all four false negative results occurred in patients whose drain amylase concentrations remained below the literature-derived 1,000 U/L detection threshold to which the SensAL was tuned. The sensor never failed to respond when amylase exceeded this threshold, confirming that the missed detections reflect a biomarker limitation rather than a technical shortcoming of the sensing platform (see supplementary patient data file for individual amylase values). Additionally, a positive sensor response was associated with an approximately 14-fold increase in the odds of a true leak event (positive likelihood ratio 14.4), with positive and negative predictive values of 88% and 90% respectively (Figure 5d), highlighting SensAL as a clinically useful rule-in test. Additional sensor performance metrics and their 95% confidence intervals (Wilson method) are summarized in Table S4 in the Supplementary Information. The two false positive results, both occurring on POD1 in patients with amylase elevations above the threshold (1,619 U/L and 2,617 U/L, respectively) in the absence of a confirmed leak, are consistent with transient enzyme release from surgical residue rather than true anastomotic leakage. Taken together, every sensor response (positive or negative) was fully consistent with the measured amylase concentration, demonstrating that SensAL performed consistently well.

**Figure 5.**
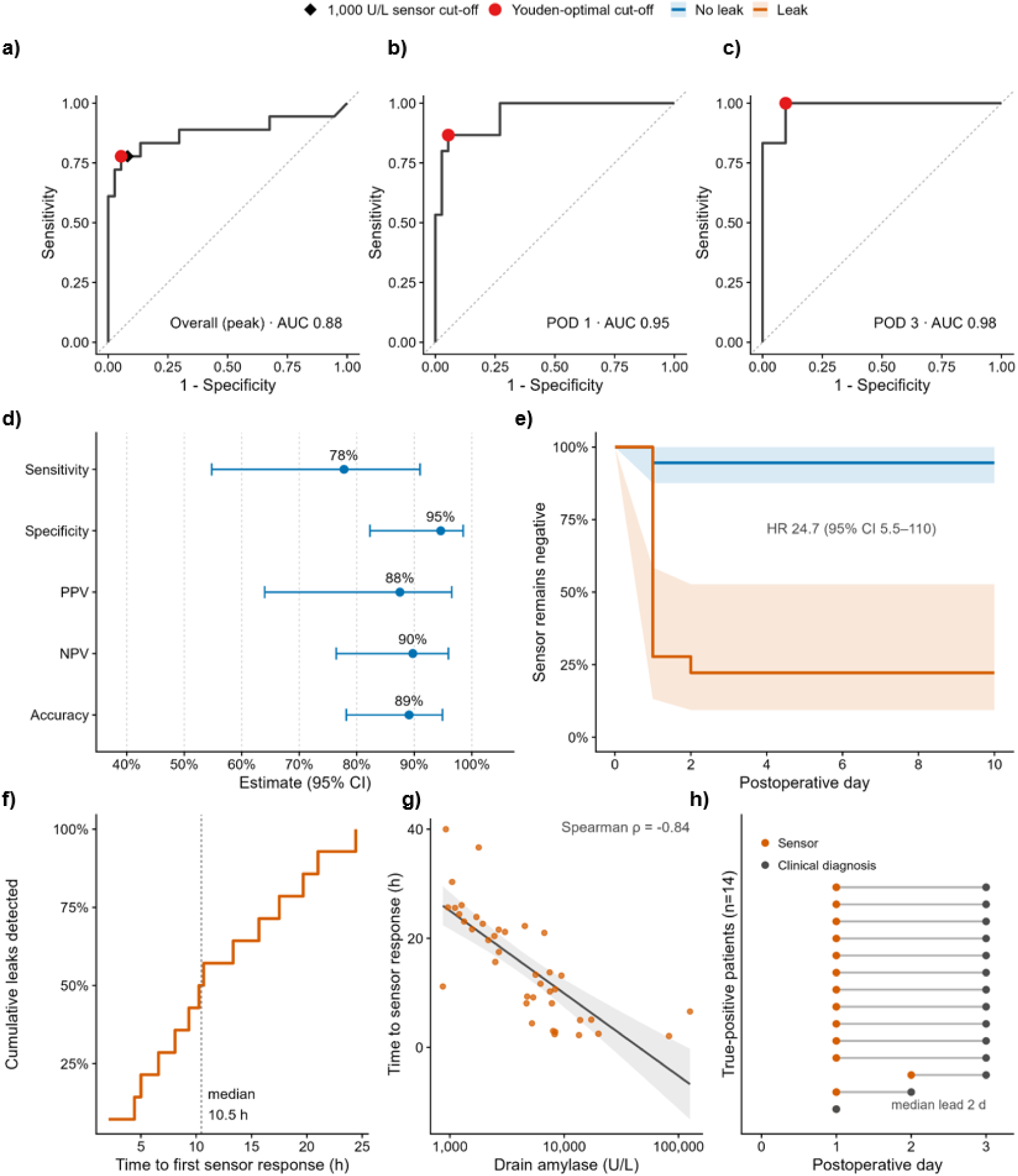
Diagnostic performance and response characteristics of the SensAL sensor in the Switzerland cohort. (a–c) Receiver-operating-characteristic (ROC) curves for drain amylase as a predictor of anastomotic leak, using the peak per-patient value a), the postoperative day 1 (POD1) value b), and the POD3 value c); areas under the curve (AUC) were 0.88 (95% CI 0.75–1.00), 0.95 (0.90–1.00) and 0.98 (0.96–1.00), respectively (DeLong intervals). The red circle marks the Youden-optimal cut-off, and in a) the black diamond marks the pre-specified 1,000 U/L sensor activation threshold. d) Diagnostic accuracy of the sensor against the clinical reference standard (primary analysis: amylase-relevant leak; n = 55, 18 leaks): sensitivity 77.8% (95% CI 54.8–91.0), specificity 94.6% (82.3–98.5), positive predictive value 87.5% (64.0–96.5), negative predictive value 89.7% (76.4–95.9) and accuracy 89.1% (78.2–94.9). Points are point estimates and horizontal bars are 95% Wilson confidence intervals. e) Kaplan–Meier estimates of the probability that the sensor remains negative over postoperative time, by leak status (blue, no leak; orange, leak; shaded bands, 95% confidence intervals). Sensor positivity occurred earlier and more frequently in leak patients (Cox hazard ratio 24.7, 95% CI 5.5–110; log-rank p < 0.001). (f) Cumulative proportion of leaks detected as a function of the in-vitro time to first sensor response (median 10.5 h; dashed line). g) Relationship between drain amylase concentration and in-vitro time to sensor response across sensor-positive leak samples (n = 40); higher drain amylase was associated with a shorter time to response (Spearman ρ = −0.84; line, log-linear fit with 95% confidence band). h) Timing of sensor positivity relative to clinical diagnosis in the 14 true-positive patients. Each row is one patient; the orange marker denotes the postoperative day of the first sensor-positive sample and the grey marker the day of clinical diagnosis (median lead 2 days). These values are at postoperative-day resolution and, because this cohort was tested in vitro on stored drain fluid against an amylase-based reference standard, are subject to incorporation and verification bias; real-time lead-time evidence is provided by the prospective Colombia cohort (Figure 5e). AUC, area under the curve; CI, confidence interval; POD, postoperative day; ROC, receiver operating characteristic.

Having confirmed that SensAL reliably detects elevated amylase, we next examined how rapidly this detection occurs. The temporal evolution of sensor responses was analyzed using Kaplan–Meier analysis (see Figure 5e) to determine when sensors transitioned from negative to positive in leak and non-leak patients. In non-leak patients, the probability of maintaining a negative sensor status remained close to 100% at POD1 and stabilized at approximately 94% thereafter. In contrast, leak patients showed a rapid decline in negative sensor probability, reaching 26% on POD1 and 22% on POD2 (Cox proportional-hazards model, hazard ratio 24.7, 95% CI 5.5–110; p < 0.001) (see also Table S5 in the Supplementary Information). These results indicate that the majority of leak events were detected by POD2, demonstrating the rapid detection capability of the sensor. To further evaluate the response kinetics of the sensing elements, the time to leak detection was analyzed across all positive sensor responses. For this the time until digestion was recorded for the first POD at which the sensor responded. The cumulative detection curve shows that leak events were progressively identified over time, with the first detections occurring 5 h after exposure to patient drain fluid. Following exposure to leak-containing fluid, the sensing elements reacted within a median of 10.5 h (range 2.1-24.4 h), with shorter times at higher amylase concentrations (Figure 5f). The corresponding amylase concentrations associated with each detection event are shown in the adjacent panel, enabling direct comparison between enzyme levels and sensor response kinetics (Figure 5g). In addition, the earlier diagnostic potential of the sensor was assessed by comparing the time of the first positive sensor response with the time of clinical leak diagnosis (Figure 5h). The sensor consistently identified leak events before clinical diagnosis, with a median lead time of 2 days, highlighting the capability of the sensing system to provide substantially earlier warning of developing leaks. Together, these results demonstrate that the enzyme-responsive sensing elements provide rapid optical detection of elevated digestive enzyme activity, enabling clinically relevant leak identification rapidly after increase of drain amylase values.

### In Clinic Sensor Performance in Hospitals in Colombia

Building on these promising results, we next deployed SensAL directly within a hospital environment in Colombia to validate its performance under real-world clinical conditions, enrolling a total of 37 patients undergoing gastrointestinal surgery in two clinics in Bucaramanga, in this prospective observational cohort study. The cohort comprised patients undergoing elective and emergency procedures in which a gastrointestinal anastomosis was performed, and a peri-anastomotic drain was placed at the surgeon’s discretion. Among the included patients, 7 (19%) were clinically diagnosed with postoperative leakage (Figure 5a). Details regarding the complete patient cohort and surgical procedures are provided in the supplementary patient data table. To replicate clinically relevant monitoring conditions, the lyophilized and sterilized sensors were integrated into fluid collection cups positioned next to the patients’ bed. Clinical personnel involved in patient care were blinded to sensor changes, whereas independent clinicians from the research team collected patient data and documented sensor responses by taking images of them. We demonstrate reliable visual detection under these conditions using TiO pigments rendering the hydrogels opaque and white, a color rarely observed in drain effluents (see selection of colors from patient drain fluids in Figure 6b), thereby enhancing contrast and facilitating straightforward visual monitoring of sensor degradation. Representative examples of reacted and unreacted sensors are shown in Figure 6c. In a representative patient who developed a leak, the amylase-responsive sensing element disappeared on POD6, while the additionally PEG crosslinked amylase-responsive element persisted until POD10, demonstrating the semiquantitative readout capability of the platform Having established the monitoring setup and confirmed sensor functionality under real-world conditions, we next evaluated the overall diagnostic performance of SensAL in this cohort. The results of this analysis are shown in Figure 6d. The performance metrics indicate strong diagnostic capability of the sensor in the clinical setting. The system achieved 100% sensitivity, correctly identifying all leak cases, and a specificity of 93%, indicating that the vast majority of non-leak patients were correctly classified. The positive predictive value was 78% (95% CI, 45-94%) reflecting two false-positive detections, whereas the negative predictive value of 100% (95% CI, 88-100%) demonstrates that patients without a sensor response were reliably leak-free. A table showing the full statistical evaluation of the sensor characteristics with 95% confidence intervals can be found in Table S6. Notably, both false-positive sensor responses occurred during later postoperative days (POD 5-6 and 7-9, respectively). Whether these cases reflect single-day threshold exceedances due to surgery residues as observed in the Swiss cohort, or a cumulative effect where prolonged exposure to mildly elevated amylase levels collectively exceeds the total enzymatic load the sensing elements can withstand, cannot be determined without concurrent amylase measurements, which will be included in future in hospital studies. This limitation, which became apparent under real-world monitoring conditions, could be easily addressed by increasing the PEGDA crosslinking density within the sensing elements, raising the total enzymatic load required to trigger a response while preserving sensitivity to the transient large enzyme elevations characteristic of true leak events. Importantly, this remains a biomarker phenomenon rather than a sensor failure, as SensAL responded to the cumulative enzymatic activity it was exposed to. Overall, the sensor reached an accuracy of 95% (95% CI, 82–99%), highlighting its potential as a reliable bedside monitoring tool. In a clinical context, the high sensitivity and negative predictive value are particularly important, as they suggest that the system can confidently rule out leaks and enable early identification of true leak events, supporting faster clinical decision-making and timely intervention. This was further corroborated by the diagnostic lead time analysis, which revealed that the sensor identified leaks a median of 5 days ahead of clinical diagnosis, with most detections occurring between 2 and 6 days earlier and the earliest detection happening 10 days prior to clinical recognition (for specific case lead time see Figure 6e). Notably, this earlier detection window could afford clinicians additional time to initiate further diagnostic evaluation or therapeutic interventions, contributing to the mitigation of severe complications associated with delayed leak recognition.

**Figure 6.**
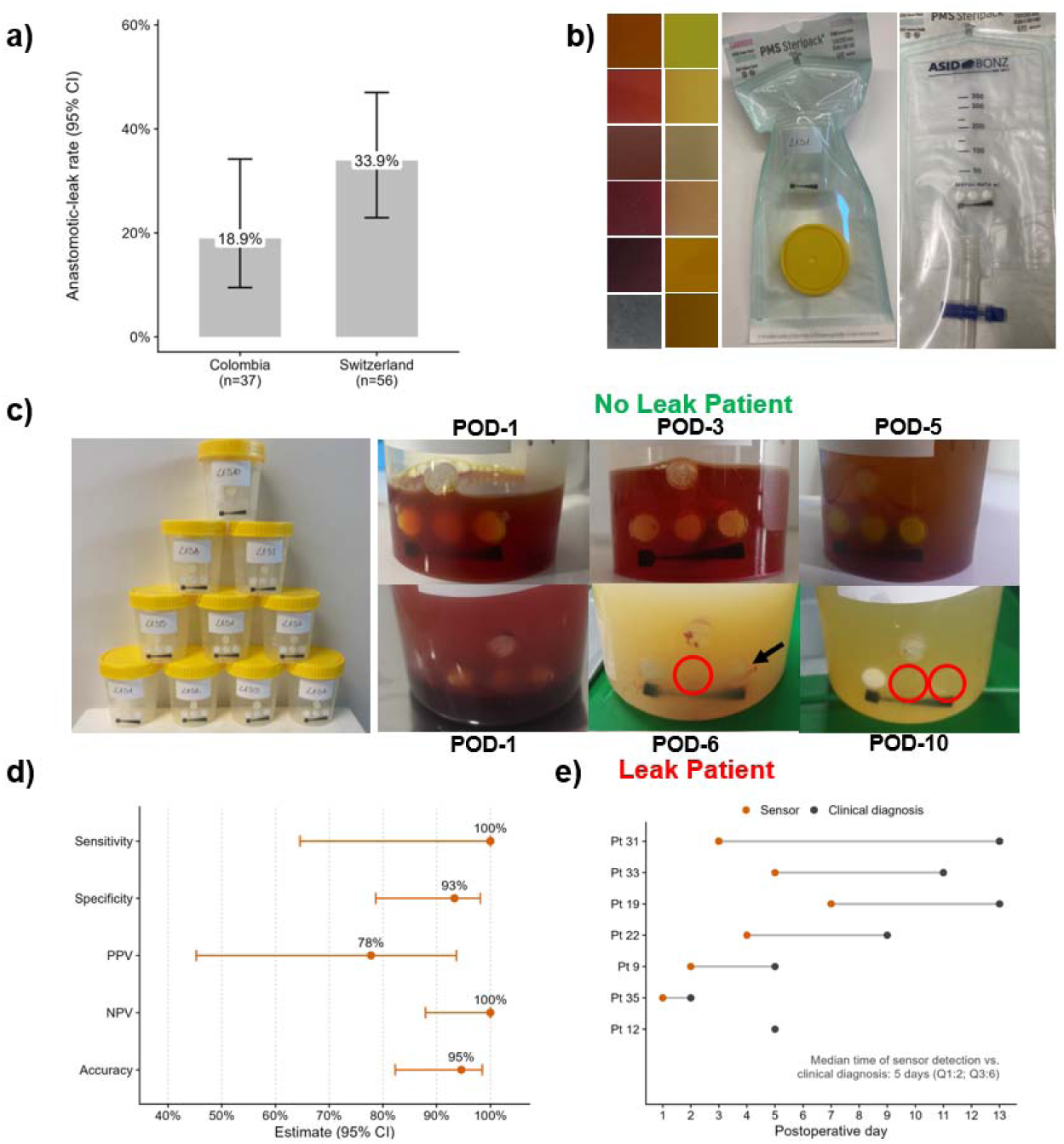
Clinical validation of the SensAL sensor in the prospective Colombia cohort. a) Anastomotic-leak rate in each study sample: Switzerland 33.9% (19/56; 95% CI 22.9–47.0) and Colombia 18.9% (7/37; 95% CI 9.5–34.2). Bars show the observed proportion; error bars are 95% Wilson confidence intervals. b) Drain fluid rainbow depicting the different drain fluid colorations encountered in routine clinical practice (photographs taken during the study in Colombia). Due to the variety of colors, TiO2 was chosen as a colorant for the study. c) Collection cups used for the clinical study in Colombia. Drain fluid from the patient’s drain was filled in these cups every 12h. In the case of a leak, the enzymes in this fluid react with the sensing elements causing them to disappear while the sensor remains unchanged in case of no leak. d) Diagnostic performance of the sensor in the Colombia cohort against the clinical reference standard (n = 37; 7 leaks): sensitivity 100% (95% CI 64.6–100), specificity 93.3% (78.7–98.2), positive predictive value 77.8% (45.3–93.7), negative predictive value 100% (87.9–100), and overall accuracy 94.6% (82.3–98.5). Points are point estimates and horizontal bars are 95% Wilson confidence intervals. e) Timing of sensor activation relative to clinical diagnosis in the seven true-positive patients. Each row is one patient; the orange marker denotes the postoperative day of sensor activation and the grey marker the day of clinical diagnosis. The sensor became positive a median of 5 days earlier than the clinical diagnosis (IQR 2–6; range 0–10; paired Wilcoxon signed-rank test, p = 0.036). CI, confidence interval; IQR, interquartile range; POD, postoperative day.

## CONCLUSIONS

In this work, we addressed one of the most feared complications in gastrointestinal surgery by introducing a low-cost, non-electronic bedside monitoring system for the early detection of anastomotic leaks. Through systematic optimization of the amylase-responsive hydrogel sensing elements, we demonstrated their potential for real-world clinical use. Importantly, the sensing elements can be lyophilized and sterilized without loss of functionality, enabling long-term storage and straightforward deployment while maintaining extremely low production costs through the use of ethylene oxide. This approach establishes a platform for simple, electronic-free, continuous, and semi-quantitative naked-eye detection of anastomotic leaks directly at the patient’s bedside. Using over 300 clinical drain fluid samples, we characterized the postoperative amylase dynamics of 56 patients during their hospital stay. The performance of the proposed sensor technology was subsequently demonstrated in a controlled validation study, where the system achieved a sensitivity of 78% and a specificity of 95%. Notably, sensor readout preceded the clinical diagnosis of leaks by a median of 2 days, highlighting the potential of the system to enable substantially earlier detection of anastomotic leaks. These results were further corroborated in a first in-hospital feasibility study, where the sterilized sensors were deployed under real-world clinical conditions to hospitals in Colombia. In this cohort of 37 patients, the sensor platform achieved 100% sensitivity and a 100% negative predictive value. These findings highlight the robustness and clinical potential of the platform for continuous bedside monitoring of anastomotic leaks, while the freeze-dried design enables long-term storage under simple conditions, facilitating practical deployment particularly in resource limited settings. Nonetheless, further studies are required to extend the present in-hospital evaluation toward a first-in-human clinical study.

On a broader scale, this low-cost sensing approach offers a promising strategy for resource-efficient identification of patients at risk of anastomotic leaks. By providing an early and easily interpretable indication of potential leak events, the system can support clinicians in deciding when to initiate additional diagnostic procedures, such as fluoroscopy, CT, or MRI, or when to adjust postoperative management strategies, including the early reintroduction of enteral nutrition. Owing to its straightforward visual readout and minimal requirement for specialized infrastructure or personnel, the sensor can be seamlessly integrated into existing hospital workflows with minimal resource consumption. Consequently, the presented sensing platform has the potential to reduce both over- and under-diagnosis of leaks while improving the efficiency of clinical decision-making and resource allocation. Ultimately, this detection strategy may contribute to improved patient outcomes while enhancing the economic sustainability of healthcare systems, particularly in low- and middle-income countries.

## EXPERIMENTAL

All materials were purchased from Sigma-Aldrich (Merck). PEG-DA was used without further purification. Human samples were received from the cantonal hospital of St. Gallen (KSSG, approved by the Cantonal Ethics Commission of the Canton of St. Gallen, Switzerland, BASEC no. Req-2022-01026). In hospital test in Colombia were approved by Universidad Industrial de Santander (Minutes No. 12, 20 June 2025), Chicamocha Clinic (Minutes No. 3, 9 July 2025), and Santander University Hospital (Minutes No. 8, 29 August 2025). Written informed consent was obtained from all participants.

### Gelatin-MA Synthesis and Hydrogel Formation

The synthesis of Gel-MA was performed as described in our previous report.^30^ For the preparation of Gel-MA hydrogel sensing elements, 5 wt.% gelatin methacrylate was dissolved in MQ water containing 1 wt.% TiO_2_ (mesh size) and heated on a magnetic stir plate at 55°C under constant stirring for 3h. Afterwards, 10 vol% Lithium phenyl-2,4,6-trimethylbenzoylphosphinate (LAP, 6.33mg/mL in MQ) was added and briefly stirred. The mixture was then pipetted on a glass plate and evenly spread with a film applicator to form a thin film of approximately 1 mm. The whole glass plate was then placed in a fridge for 15 minutes and the plate was irradiated with UV light (UVASPOT 400/T mercury lamp, Hönle) for 5 minutes at a distance of 15 cm. Next, a 4 mm biopsy punch was used to stencil out circular elements, which were then kept in a falcon tube in the fridge for several days until further use.

### Starch-Glycidylacrylate Synthesis and Hydrogel Formation

Starch-GMA was synthesized using a modified version of the protocol of Reis et.al..^32^ In short a 100 mL round bottom was equipped with a magnetic stir bar and 60 mL DMSO. Stirring was initiated and 4g Starch (from potatoes) and 1.5g 4-(Dimethylamino)pyridin (DMAP) was added. The mixture was then stirred at RT for 20 minutes at 100 rpm. Next 1.7 mL of Gylcidyl methacrylate (GMA, with 100 ppm monomethyl ether hydroquinone as inhibitor) was added dropwise with a syringe. The reaction was then stirred at 100 rpm for 24 hours after which 50 mL MQ was added and stirring was continued for 30 minutes. Next the mixture was transferred into dialysis tubing (3.5 kDa cutoff, Servapor) and dialyzed against 10 L deionized water. Water was exchanged after 24 hours and dialysis was continued for an additional 3 days. Next the dialyzed solution was transferred into falcon tubes and frozen at -20°C overnight. The frozen Starch-GMA was then freeze dried for 6 days and stored in a freezer until it was used. To produce the starch hydrogel sensing elements, 10 wt.% starch was suspended in 1 wt.% TiO_2_ (mesh size) suspension in MQ and vortexed for 2 minutes. The hydrogel pre.mix was then homogenized with a homogenizer (which) until no visible chunks were left. The mix was then heated at 85°C for 30 minutes, afterwards briefly vortexed and then stirred on a heating plate at 55°C and 300 rpm for 3 hours. The mixture was then pipetted on a glass plate and evenly spread with a film applicator to form a thin film of approximately 1 mm. The whole glass plate was then placed in a fridge for 15 minutes and the plate was irradiated with UV light (UVASPOT 400/T mercury lamp, Hönle) for 5 minutes at a distance of 15 cm. Next, a 4 mm biopsy punch was used to stencil out circular elements, which were then kept in a falcon tube in the fridge for several days until further use.

### Hydrogel Sensor Swelling Study

The procedure used to characterize the swelling behavior of the three hydrogels, GelMA, Starch-GMA and Starch-GMA+PEGDA, was adapted from protocols based on H. Na et al.^37^ and H. Hezaveh et al.^38^ Experiments were done in triplicates and results are reported as mean values of the individual measurements, with error bars indicating standard deviation. Each sensing element (prepared as previously mentioned) was immersed in a 50 mL Falcon Tube filled with 30mL of Dulbecco’s Phosphate Buffered Saline (DPBS) modified with MgCl_2_ and CaCl_2_. Between measurements, the tube was placed on a moving plate between the measurements. For accurate mass determination, sensing elements were removed from the PBS at regular time intervals and briefly placed on weighing paper on the scale to measure their mass. After returning the sensing element into the same centrifuge tube, any residual PBS remaining on the weighing paper was weighed separately and subtracted from the measured value. The swelling percentage was calculated according to the following formula, where m(0) represents the mass of the dry gel at time 0 and m(t) denotes the mass of the swollen sensing element at time t:

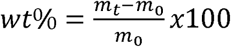

### Rheology

Sensing elements were prepared as previously mentioned. For the rheology measurements, a 8mm biopsy punch was used to make the sensing elements in order to fit the dimensions of the rheometer (Anton Paar, MCR102e). Freshly prepared SE as well as lyophilized and sterilized SE were pre-swollen for 24h in MQ water before measurement. Each SE type was measured in triplicates and results shown are average values of these measurements.

### Sensing Element Digestion Kinetics

The 4 mm sensing elements used for these experiments were produced as previously mentioned. The SE were then placed in a 48 well plate and 100 µL of the corresponding enzyme solution in DPBS modified with MgCl_2_ and CaCl_2_ was added to the well. Sterilized and lyophilized hydrogels were swollen in the same medium for 24h prior to exposure to enzymes. The plate was then mounted on a 3D printed scaffold which was placed on an orbital shaker set at 180rpm. Beneath the scaffold, a camera (GoPro Hero12) was placed and a picture of the SE in enzyme solution was taken every 5 minutes for 36 hours at room temperature. The pictures were then manually evaluated and the time point at which the sensing element disappeared was noted.

### Clinical Sample Testing (Switzerland)

Samples were collected from Kantonsspital St. Gallen (HOCH-Health Ostschweiz) with an ethics approval in place (BASEC no. Req-2022-01026). Volumes, colors as well as consistencies of the samples varied. Type of operation/complication and date collected was noted in anonymized form and the study was performed in a blinded setting (clinician holding the key). If possible, samples were collected each postoperative day in 50 mL Falcon Tubes filled with 1000 I.E. Heparin (Liquemin) by dedicated study personnel and stored at -20°C until testing. Testing was conducted in the same manner as for the kinetics measurements described above. From each sample tested approx. 5 mL was placed in a heparin coated 10 mL blood collection container (BD Vacutainer, LH 170 I.U) and refrozen until analysis by the Institute for Clinical Chemistry (IKC) of USZ, where amylase and lipase activity of the samples were determined.

### In Hospital Testing (Colombia)

The hydrogel formulation and preparation method used for the in hospital study is the same as described above. In this case the hydrogel mix was filled into a 3D printed backing with a syringe and after polymerization the backings were frozen overnight. After lyophilization, the backings were glued into the fluid collection cups and the sensor containing cups were then packed into Tyvek pouches and sterilized by ethylene oxide (Andersen, Anprolene AN75). After shipping the sensors to the hospital, they were stored at room temperature until use.

The cups were placed in a sealable, non-transparent container at the patient’s bedside. Every 12 h after surgery, drain fluid from the patient’s surgical drainage bag was transferred into the cups and a photograph of the sensor was taken. Sensors were kept in the collection cups for the entire duration of the patients’ hospital stay and were not replaced during the monitoring period. The attending doctors were not involved in the evaluation of the sensor readout and remained blinded throughout the patient’s hospital stay. After patient discharge, the sensor’s daily readout was compared with the clinical diagnosis, established through standard-of-care procedures including the patient’s clinical status, routine serum tests, imaging findings, or intraoperative findings. In no case was the clinical diagnosis based on amylase concentration in peritoneal fluid, as this parameter is not part of routine postoperative assessment at the participating centers. For the statistical evaluation, a container containing at least one fully reacted sensing element in a patient with a clinically confirmed leak was classified as a true positive event, and a confusion matrix was constructed from the recorded sensor outcomes and clinical diagnoses to calculate diagnostic performance metrics.

### Statistics

#### Diagnostic accuracy

Sensitivity, specificity, positive and negative predictive value, and overall accuracy were derived from the 2×2 classification of sensor response against the reference standard, with 95% Wilson confidence intervals; positive and negative likelihood ratios and the diagnostic odds ratio were obtained with a Haldane correction for zero cells, and combinations containing a structural zero (e.g., no false negatives) are reported as not estimable. The discriminative ability of drain amylase for leak was characterized by receiver-operating-characteristic analysis (maximum per-patient, POD1, and POD3 concentrations), with areas under the curve and both DeLong and 2,000-replicate bootstrap 95% confidence intervals; the pre-specified 1,000 U/L operating point and the Youden-optimal threshold are reported.

#### Postoperative amylase dynamics

For each patient and postoperative day (POD), the peak drain amylase was modelled on the log scale using a linear mixed-effects model with fixed effects for leak status, POD (POD1– 7, as a factor), and their interaction, and a patient-level random intercept to account for the within-patient correlation of repeated measurements; approximately half of the total variance in amylase was attributable to between-patient differences (intraclass correlation 0.50), confirming the need to model this clustering.

The group×time interaction and the overall group effect were tested by likelihood-ratio tests against nested models. Group-by-POD geometric means and leak/non-leak fold-changes with 95% confidence intervals were obtained from estimated marginal means and back-transformed from the log scale, and the intraclass correlation was derived from the model’s variance components. Two pre-specified sensitivity analyses were performed: a left-censored (Tobit) model accommodating the 10 U/L assay floor, and per-POD Wilcoxon rank-sum tests with Benjamini–Hochberg control of the false-discovery rate across POD1-7, summarized by the Hodges-Lehmann shift (fold-change) and the rank-biserial correlation.

#### Timing and lead time

Time from surgery (POD) to first sensor positivity was compared between leak and non-leak patients using Kaplan-Meier estimation, the log-rank test, and Cox proportional-hazards regression (Efron handling of ties); patients who never became positive were censored at their last sampled POD. Analytical (invitro) response time was summarized descriptively. Diagnostic earliness was quantified at day resolution among true-positive patients as the interval between clinical diagnosis and first sensor positivity; in Colombia, sensor and clinical diagnosis days were compared by the paired Wilcoxon signed-rank test.

#### Software

Analyses were performed in Python, Origin, and R 4.5.1 using lme4, lmerTest, emmeans, survival, pROC, DescTools, epiR, and cutpointr. All tests were two-sided with α = 0.05, and all confidence intervals are 95%.

## Supporting information

Patient Data

## Data Availability

All data produced in the present study are available upon reasonable request to the authors

## AUTHOR CONTRIBUTIONS

I.K.H. conceived the project idea. A.J. developed the macromolecular sensing elements, planned and executed experiments and wrote the manuscript. I.F. did the statistical evaluation of the clinical data. C.H. conducted the testing of the patient samples. S.A.G. helped with the statistical analysis of the clinical data. A.W. conducted the swelling experiments. F.S. designed and created the sensor backing. J.C.H, D.C.Q, J.M.P, A.C.Q and J.P.S. planned and executed the in-hospital study in Colombia. A.S. provided clinical insight, I.T. planned and executed the sample collection in St. Gallen. All authors reviewed, contributed, and approved the final version of the manuscript.

## ACKNOWLEDGEMENTS

We kindly acknowledge funding from the Swiss National Science Foundation (Eccellenza grant no. 181290 and Project grant no. IC00I0L-227785 to I.K.H.), the Evi Diethelm-Winteler-Stiftung (I.K.H.) the ETH Domain Specific Focus Area Personalized Health and Related Technologies (PHRT) program (grant number 2023/982 to I.K.H.) and the Swiss National Science Foundation BRIDGE Discovery Program (grant number 40B2-0_226647 to I.K.H.). The Colombian trial was supported by the Leading House for Latin America through a Research Partnership Grant.

## REFERENCES

1. Rose, J. et al. Estimated need for surgery worldwide based on prevalence of diseases: a modelling strategy for the WHO Global Health Estimate. The Lancet Global Health 3, S13–S20 (2015).

2. Goulder, F. Bowel anastomoses: The theory, the practice and the evidence base. World J Gastrointest Surg 4, 208–213 (2012).

3. Ortigão, R. et al. Anastomotic Leaks following Esophagectomy for Esophageal and Gastroesophageal Junction Cancer: The Key Is the Multidisciplinary Management. GE - Portuguese Journal of Gastroenterology 30, 38–48 (2023).

4. Hosotani, R., Doi, R. & Imamura, M. Duct-to-mucosa Pancreaticojejunostomy Reduces the Risk of Pancreatic Leakage after Pancreatoduodenectomy. World Journal of Surgery 26, 99–104 (2002).

5. Ellis, C. T. & Maykel, J. A. Defining Anastomotic Leak and the Clinical Relevance of Leaks. Clinics in Colon and Rectal Surgery 34, 359–365 (2021).

6. Xu, H. & Kong, F. Malnutrition-Related Factors Increased the Risk of Anastomotic Leak for Rectal Cancer Patients Undergoing Surgery. Biomed Res Int 2020, 5059670 (2020).

7. Zarnescu, E. C., Zarnescu, N. O. & Costea, R. Updates of Risk Factors for Anastomotic Leakage after Colorectal Surgery. Diagnostics (Basel) 11, 2382 (2021).

8. Perera, S. K. et al. Global demand for cancer surgery and an estimate of the optimal surgical and anaesthesia workforce between 2018 and 2040: a population-based modelling study. The Lancet Oncology 22, 182– 189 (2021).

9. Pedrazzoli, S. Pancreatoduodenectomy (PD) and postoperative pancreatic fistula (POPF): A systematic review and analysis of the POPF-related mortality rate in 60,739 patients retrieved from the English literature published between 1990 and 2015. Medicine 96, e6858 (2017).

10. Bassi, C. et al. The 2016 update of the International Study Group (ISGPS) definition and grading of postoperative pancreatic fistula: 11 Years After. Surgery 161, 584–591 (2017).

11. DeOliveira, M. L. et al. Assessment of Complications After Pancreatic Surgery: A Novel Grading System Applied to 633 Patients Undergoing Pancreaticoduodenectomy. Annals of Surgery 244, 931 (2006).

12. Predictors of septic shock following anastomotic leak after major gastrointestinal surgery: An audit from a tertiary care institute. Indian Journal of Critical Care Medicine 17, 298–303 (2013).

13. Wente, M. N. et al. Postpancreatectomy hemorrhage (PPH)–An International Study Group of Pancreatic Surgery (ISGPS) definition. Surgery 142, 20–25 (2007).

14. Komen, N., Bruin, R. W. F. D., Kleinrensink, G. J., Jeekel, J. & Lange, J. F. Anastomotic leakage, the search for a reliable biomarker. A review of the literature. Colorectal Disease 10, 109–115 (2008).

15. Almeida, A. B. et al. Elevated serum C-reactive protein as a predictive factor for anastomotic leakage in colorectal surgery. International Journal of Surgery 10, 87–91 (2012).

16. Lee, J. M., Lee, J., Kim, T. & Kim, N. K. Early Detection of Anastomotic Leak via the Drain/Serum Amylase Ratio in Patients Undergoing Colorectal Surgery, Particularly in Ileal Anastomosis. Yonsei Med J 66, 482–490 (2025).

17. Messias, B. A. et al. Serum C-reactive protein is a useful marker to exclude anastomotic leakage after colorectal surgery. Scientific reports 10, 1–8 (2020).

18. McKechnie, T. et al. Using preoperative C-reactive protein levels to predict anastomotic leaks and other complications after elective colorectal surgery: A systematic review and meta-analysis. Colorectal Disease 26, 1114–1130 (2024).

19. Towe, C. et al. Drain Amylase Can Accurately Detect Anastomotic Leak Independent of Patient Factors and Location or Type of Anastomosis. Journal of the American College of Surgeons 229, e56 (2019).

20. Linden, P. A. et al. Drain Amylase: A Simple and Versatile Method of Detecting Esophageal Anastomotic Leaks. The Annals of Thoracic Surgery 113, 1794–1800 (2022).

21. Berkelmans, G. H. K. et al. Diagnostic value of drain amylase for detecting intrathoracic leakage after esophagectomy. World J Gastroenterol 21, 9118–9125 (2015).

22. Amroun, K. et al. High amylase concentration in drainage liquid can early predict proximal and distal intestinal anastomotic leakages: A prospective observational study. J Res Med Sci 28, 5 (2023).

23. Clark, D. A. et al. Drain fluid amylase as a biomarker for the detection of anastomotic leakage after rectal resection without a diverting ileostomy. ANZ Journal of Surgery 92, 813–818 (2022).

24. Schots, J. P. M., Luyer, M. D. P. & Nieuwenhuijzen, G. A. P. Abdominal Drainage and Amylase Measurement for Detection of Leakage After Gastrectomy for Gastric Cancer. Journal of Gastrointestinal Surgery 22, 1163–1170 (2018).

25. Ben-David, M. et al. Implantation of an Impedance Sensor for Early Detection of Gastrointestinal Anastomotic Leaks. Journal of Surgical Research 278, 49–56 (2022).

26. Huynh, M. et al. Continuous pH monitoring using a sensor for the early detection of anastomotic leaks. Front. Med. Technol. 5, (2023).

27. Komen, N., De Bruin, R. W. F., Kleinrensink, G. J., Jeekel, J. & Lange, J. F. Anastomotic leakage, the search for a reliable biomarker. A review of the literature. Colorectal Disease 10, 109–115 (2008).

28. Walshaw, J., Hugh, K., Helliwell, J., Burke, J. & Jayne, D. Perianastomotic pH Monitoring for Early Detection of Anastomotic Leaks in Gastrointestinal Surgery: A Systematic Review of the Literature. Surg Innov 32, 180–195 (2025).

29. ShafieiDarabi, M. et al. Microwave biosensor for amylase detection in drainage fluid to monitor anastomotic leakage. Biosensors and Bioelectronics 290, 117990 (2025).

30. Jessernig, A. et al. Early Detection and Monitoring of Anastomotic Leaks via Naked Eye-Readable, Non-Electronic Macromolecular Network Sensors. Advanced Science 11, 2400673 (2024).

31. Kunyanee, K. et al. Improving the swelling capacity of granular cold-water rice starch by ultrasound-assisted alcoholic-alkaline treatment. Ultrason Sonochem 98, 106506 (2023).

32. Reis, A. V. et al. Synthesis and characterization of a starch-modified hydrogel as potential carrier for drug delivery system. Journal of Polymer Science Part A: Polymer Chemistry 46, 2567–2574 (2008).

33. Teixeira, U. F. et al. EARLY DRAIN FLUID AMYLASE IS USEFUL TO PREDICT PANCREATIC FISTULA AFTER PANCREATODUODENECTOMY: LESSONS LEARNED FROM A SOUTHERN BRAZILIAN CENTER. Arq. Gastroenterol. 55, 160–163 (2018).

34. van Dongen, J. C., Merkens, S., Aziz, M. H., Groot Koerkamp, B. & van Eijck, C. H. J. The value of serum amylase and drain fluid amylase to predict postoperative pancreatic fistula after pancreatoduodenectomy: a retrospective cohort study. Langenbecks Arch Surg 406, 2333–2341 (2021).

35. Liu, Y., Li, Y., Wang, L. & Peng, C.-J. Predictive value of drain pancreatic amylase concentration for postoperative pancreatic fistula on postoperative day 1 after pancreatic resection: An updated meta-analysis. Medicine 97, e12487 (2018).

36. Wiedemann, D. et al. Biochemical Early Detection of Postoperative Pancreatic Fistula. Visc Med 41, 121– 129 (2025).

37. Na, H. et al. Hydrogel-based strong and fast actuators by electroosmotic turgor pressure. Science 376, 301– 307 (2022).

38. Hezaveh, H. & Muhamad, I. I. Modification and swelling kinetic study of kappa-carrageenan-based hydrogel for controlled release study. Journal of the Taiwan Institute of Chemical Engineers 44, 182–191 (2013).

