## Supplementary Information for "Sterilizable, Time-Integrating Hydrogel Sensors Enable Continuous Gastrointestinal Leak Surveillance in Low- and High-Resource Settings"

**Supplementary Tables**

Table S1. Drain amylase by postoperative day — leak patients (per-patient peak; amylase-relevant leak, n=18).

| **POD** | **n** | **Median (IQR), U/L** | **Model geom. mean (95% CI), U/L** | **Range, U/L** | **>1000 U/L, %** | **At floor (≤10 U/L), %** |
| --- | --- | --- | --- | --- | --- | --- |
| 1 | 15 | 4,675 (2,056–7,639) | 3,172 (1,675–6,007) | 461–126,000 | 86.7 | 0.0 |
| 2 | 14 | 1,300 (821–5,105) | 1,448 (753–2,785) | 183–17,276 | 57.1 | 0.0 |
| 3 | 12 | 576 (282–1,120) | 513 (258–1,022) | 92–4,526 | 25.0 | 0.0 |
| 4 | 15 | 128 (54–436) | 139 (73–263) | 13–7,517 | 6.7 | 0.0 |
| 5 | 12 | 106 (26–247) | 114 (57–226) | 10–8,004 | 16.7 | 8.3 |
| 6 | 10 | 164 (16–966) | 162 (78–337) | 10–9,456 | 30.0 | 10.0 |
| 7 | 7 | 51 (22–1,928) | 194 (84–450) | 10–8,348 | 28.6 | 28.6 |
| 8 | 4 | 2,124 (1,176–5,382) | — | 10–13,483 | 75.0 | 25.0 |
| 9 | 2 | 41,330 (20,670–61,989) | — | 10–82,649 | 50.0 | 50.0 |
| 10 | 2 | 9,978 (4,994–14,963) | — | 10–19,947 | 50.0 | 50.0 |
| IQR, interquartile range. Median (IQR) and range are descriptive statistics of the observed per-patient peak values. | | | | | | |
| Model geom. mean (95% CI) is the back-transformed estimated marginal mean from the linear mixed-effects model (fitted to POD1–7) and is the value reported in the main text; '—', beyond the modelled range. | | | | | | |

Table S2. Drain amylase by postoperative day — no-leak patients (n=37).

| **POD** | **n** | **Median (IQR), U/L** | **Model geom. mean (95% CI), U/L** | **Range, U/L** | **>1000 U/L, %** | **At floor (≤10 U/L), %** |
| --- | --- | --- | --- | --- | --- | --- |
| 1 | 37 | 95 (41–488) | 115 (76–175) | 10–2,617 | 8.1 | 8.1 |
| 2 | 30 | 44 (17–113) | 57 (36–89) | 10–829 | 0.0 | 20.0 |
| 3 | 31 | 27 (10–50) | 28 (18–44) | 10–177 | 0.0 | 25.8 |
| 4 | 34 | 14 (10–27) | 18 (11–27) | 10–66 | 0.0 | 41.2 |
| 5 | 28 | 12 (10–22) | 16 (10–25) | 10–45 | 0.0 | 42.9 |
| 6 | 24 | 10 (10–20) | 16 (10–26) | 10–118 | 0.0 | 54.2 |
| 7 | 19 | 12 (10–22) | 16 (10–28) | 10–114 | 0.0 | 42.1 |
| 8 | 12 | 12 (10–30) | — | 10–65 | 0.0 | 33.3 |
| 9 | 8 | 11 (10–47) | — | 10–695 | 0.0 | 37.5 |
| 10 | 5 | 12 (10–35) | — | 10–63 | 0.0 | 40.0 |
| A substantial proportion of no-leak values lie at the assay floor (≤10 U/L), consistent with left-censoring. | | | | | | |
| Model geom. mean (95% CI) is the back-transformed estimated marginal mean from the linear mixed-effects model (POD1–7), as reported in the main text. | | | | | | |

Table S3. Between-group comparison of drain amylase by postoperative day.

| **POD** | **n (leak / no-leak)** | **Fold-change Leak/No-leak (95% CI)*** | **Rank-biserial r** | **Wilcoxon p** | **BH-adjusted q** | **Censored (Tobit) fold-change (95% CI)†** |
| --- | --- | --- | --- | --- | --- | --- |
| 1 | 15 / 37 | 31.3 (11.6–95.4) | 0.91 | <0.001 | <0.001 | 36.5 (14.2–93.9) |
| 2 | 14 / 30 | 36.0 (15.5–84.3) | 0.95 | <0.001 | <0.001 | 39.5 (16.0–97.5) |
| 3 | 12 / 31 | 19.1 (9.4–50.3) | 0.97 | <0.001 | <0.001 | 24.7 (11.5–53.2) |
| 4 | 15 / 34 | 7.8 (3.7–16.8) | 0.88 | <0.001 | <0.001 | 14.0 (6.3–31.0) |
| 5 | 12 / 28 | 6.2 (2.5–15.5) | 0.77 | <0.001 | <0.001 | 11.9 (4.5–32.0) |
| 6 | 10 / 24 | 2.9 (1.2–53.9) | 0.60 | 0.005 | 0.006 | 27.5 (5.2–144.7) |
| 7 | 7 / 19 | 4.3 (1.0–138.0) | 0.50 | 0.053 | 0.053 | 15.3 (2.4–98.0) |
| * Hodges-Lehmann fold-change (Leak vs No leak) from the Wilcoxon rank-sum test; BH = Benjamini-Hochberg across POD1–7. | | | | | | |
| † Sensitivity analysis using a left-censored (Tobit) model with the assay floor at 10 U/L. POD7 q=0.053; all other PODs q<0.01. | | | | | | |
| Main-text fold-changes are model-based ratios of estimated marginal means from the linear mixed-effects model (e.g., POD1 27.5, 95% CI 12.8–59.0); the Hodges-Lehmann and Tobit estimates above are method-specific corroborations and differ slightly. | | | | | | |

Table S4. Diagnostic performance of SensAL — Switzerland (in-vitro validation).

| **Metric** | **Primary: amylase-relevant leak (N=55)** |
| --- | --- |
| Sensitivity, % (95% CI) | 77.8 (54.8–91.0) |
| Specificity, % (95% CI) | 94.6 (82.3–98.5) |
| PPV, % (95% CI) | 87.5 (64.0–96.5) |
| NPV, % (95% CI) | 89.7 (76.4–95.9) |
| Accuracy, % (95% CI) | 89.1 (78.2–94.9) |
| LR+ (95% CI) | 14.4 (3.7–56.6) |
| LR− (95% CI) | 0.235 (0.099–0.559) |
| Diagnostic OR (95% CI) | 61.2 (10.1–373.1) |
| Confusion (TP/FP/FN/TN) | 14/2/4/35 |

Table S5. Time to SensAL positivity — Kaplan-Meier probability of remaining sensor-negative (Switzerland).

| **POD** | **No leak — sensor negative (n at risk)** | **Leak — sensor negative (n at risk)** |
| --- | --- | --- |
| 0 | 100.0% (n=37) | 100.0% (n=18) |
| 1 | 94.6% (n=37) | 27.8% (n=18) |
| 2 | 94.6% (n=35) | 22.2% (n=5) |
| 3 | 94.6% (n=34) | 22.2% (n=4) |
| 4 | 94.6% (n=34) | 22.2% (n=4) |
| 5 | 94.6% (n=28) | 22.2% (n=4) |
| 6 | 94.6% (n=25) | 22.2% (n=3) |
| 7 | 94.6% (n=19) | 22.2% (n=2) |
| 8 | 94.6% (n=12) | 22.2% (n=0) |
| 9 | 94.6% (n=8) | 22.2% (n=0) |
| 10 | 94.6% (n=5) | 22.2% (n=0) |
| Cox proportional-hazards HR (Leak vs No leak) = 24.7 (95% CI 5.5–110), p<0.001 (Efron ties); log-rank p<0.001. | | |

Table S6. Diagnostic performance of SensAL — Colombia (prospective in-hospital).

| **Metric** | **Primary (N=37)** |
| --- | --- |
| Sensitivity, % (95% CI) | 100.0 (64.6–100.0) |
| Specificity, % (95% CI) | 93.3 (78.7–98.2) |
| PPV, % (95% CI) | 77.8 (45.3–93.7) |
| NPV, % (95% CI) | 100.0 (87.9–100.0) |
| Accuracy, % (95% CI) | 94.6 (82.3–98.5) |
| LR+ (95% CI) | 15.0 (3.9–57.2) |
| LR− (95% CI) | Not estimable (0 FN) |
| Diagnostic OR (95% CI) | Not estimable (0 FN) |
| Confusion (TP/FP/FN/TN) | 7/2/0/28 |
